# Blood-based DNA methylation study of alcohol consumption

**DOI:** 10.1101/2024.02.26.24303397

**Authors:** Elena Bernabeu, Aleksandra D Chybowska, Jacob K. Kresovich, Matthew Suderman, Daniel L McCartney, Robert F Hillary, Janie Corley, Maria Del C. Valdés-Hernández, Susana Muñoz Maniega, Mark E. Bastin, Joanna M. Wardlaw, Zongli Xu, Dale P. Sandler, Archie Campbell, Sarah E Harris, Andrew M McIntosh, Jack A. Taylor, Paul Yousefi, Simon R Cox, Kathryn L Evans, Matthew R Robinson, Catalina A Vallejos, Riccardo E Marioni

**Affiliations:** Centre for Genomic and Experimental Medicine, Institute of Genetics and Cancer, University of Edinburgh, Edinburgh, UK; Department of Cancer Epidemiology, H. Lee Moffitt Cancer Center and Research Institute, Tampa, FL, USA; Medical Research Council Integrative Epidemiology Unit, University of Bristol, Bristol BS8 1TH, UK; Usher Institute, Edinburgh Medical School, University of Edinburgh, Edinburgh, UK; Lothian Birth Cohorts, Department of Psychology, University of Edinburgh, Edinburgh, UK; Scottish Imaging Network, A Platform for Scientific Excellence (SINAPSE) Collaboration, Edinburgh, UK; Centre for Clinical Brain Sciences, Edinburgh Imaging and UK Dementia Research Institute, University of Edinburgh, UK; Epidemiology Branch, National Institute of Environmental Health Sciences, Research Triangle Park, NC, USA; Neurovascular Imaging Research Core, UCLA, Los Angeles, CA; Division of Psychiatry, University of Edinburgh, Royal Edinburgh Hospital, Edinburgh, UK; Institute of Science and Technology Austria, Klosterneuburg, Austria; Medical Research Council Human Genetics Unit, Institute of Genetics and Cancer, University of Edinburgh, Edinburgh, UK; The Alan Turing Institute, London, UK

## Abstract

Alcohol consumption is an important risk factor for multiple diseases. It is typically assessed via self-report, which is open to measurement error and bias. Instead, molecular data such as blood-based DNA methylation (DNAm) could be used to derive a more objective measure of alcohol consumption by incorporating information from cytosine-phosphate-guanine (CpG) sites known to be linked to the trait. Here, we explore the epigenetic architecture of self-reported weekly units of alcohol consumption in the Generation Scotland study. We first create a blood-based epigenetic score (EpiScore) of alcohol consumption using elastic net penalised linear regression. We explore the effect of pre-filtering for CpG features ahead of elastic net, as well as differential patterns by sex and by units consumed in the last week relative to an average week. The final EpiScore was trained on 16,717 individuals and tested in four external cohorts: the Lothian Birth Cohorts (LBC) of 1921 and 1936, the Sister Study, and the Avon Longitudinal Study of Parents and Children (total N across studies > 10,000). The maximum Pearson correlation between the EpiScore and self-reported alcohol consumption within cohort ranged from 0.41 to 0.53. In LBC1936, higher EpiScore levels had significant associations with poorer global brain imaging metrics, whereas self-reported alcohol consumption did not. Finally, we identified two novel CpG loci via a Bayesian penalized regression epigenome-wide association study (EWAS) of alcohol consumption. Together, these findings show how DNAm can objectively characterize patterns of alcohol consumption that associate with brain health, unlike self-reported estimates.

## Introduction

Alcohol consumption, particularly heavy use, has been associated with increased morbidity and mortality, cognitive impairment, progressive white matter degeneration in the brain, and is a major risk factor for various forms of cancer^1–5^. Further, alcohol misuse can lead to alcohol use disorders (AUD) and alcohol-related organ damage^3,4,6–8^. Alcohol consumption also associates with differential DNA methylation (DNAm) patterns^9–11^; DNAm is an epigenetic mark that is typically characterized by the addition of a methyl group to the 5’ carbon of a cytosine base, often occurring at cytosine-phosphate-guanine (CpG) dinucleotides, also referred to as a CpG site^12^. DNAm can influence gene expression and cellular function, thus methylomic modifications could mediate alcohol-disease risk associations and development^12,13^ as well as alcohol addiction^6,14^. As such, identification of alcohol-associated CpG sites could provide biological insights into the pathophysiology of alcohol-related diseases^11,15^.

Despite alcohol misuse being an important risk factor for a plethora of diseases, self-reported consumption is an imperfect phenotype that can be prone to error, particularly amongst heavy drinkers^16^. An AUD diagnosis is also not a good proxy for increased consumption as most people with an alcohol-attributable disease or injury are not diagnosed^4^. Several biochemical markers exist to quantify heavy alcohol use, including serum alanine transaminase (ALT) and aspartate transaminase (AST) levels, as well as AT-rich interactive domain-containing protein 4B (ARID4B), phosphatidylcholine-sterol acyltransferase (LCAT), hepatocyte growth factor-like protein (MST1) and ADP-ribosylation factor 6 (ARL6). Together, these biomarkers have been estimated to have good but not perfect discriminatory ability of heavy alcohol consumption, as measured by an area under the receiver operating characteristic curve (AUC) of 0.73-0.86^17–19^. DNAm-based predictors may improve upon these estimates. They have gained prominence in recent years through the prediction of phenotypes such as age and smoking^11,20–22^. The previous largest epigenome wide association (EWAS) meta-analysis study of alcohol consumption (self-reported units consumed per day in the past year) included over 13,000 individuals from 13 cohorts. Using a 144-CpG signature, the authors explained up to 13.8% of the variance in the phenotype (incremental R^2^ over linear regression models including age and sex) in four independent test sets.

In this study, we explore the creation of an epigenetic predictor of alcohol consumption, making use of a large single-cohort DNAm study, Generation Scotland. We assess the performance of this predictor in 9 independent external subsets from four different studies, the Lothian Birth Cohorts (LBC) of 1921 and 1936^23,24^, the Avon Longitudinal Study of Parents and Children (ALSPAC)^25,26^, and the Sister Study^27^, and explore differential patterns by sex and units consumed in the last week relative to an average week. Furthermore, to gain further biological insights into potential alcohol-mediated pathways underlying disease, we perform the largest epigenome-wide association study (EWAS) of alcohol consumption to date (N = 16,717).

## Results

### The Generation Scotland Cohort

Generation Scotland is a Scottish family-based study with over 24,000 participants recruited between 2006 and 2011^28^. Participants were aged between 18 and 99 years at recruitment, with a mean age of 47.5 years (SD 14.9). After exclusions (**Supplementary Figure 1**), a total of 16,717 participants (9,758 females and 6,959 males) had measured blood-based DNAm (see **Methods**) and self-reported alcohol consumption data available (**Supplementary Table 1 and 2**). The mean units consumed in the week prior to completing the questionnaire and blood draw was 10.9 (SD 12.7, **Supplementary Figure 2 and 3**). A total of 10,506 (62.8%) participants reported that this number was reflective of their usual drinking pattern with 1,622 and 3,756 noting it was less or more than they typically drink in a week (response unknown for N = 833).

### Alcohol consumption EpiScore – filtering by input features and trait definition

An epigenetic score (EpiScore) was trained on self-reported alcohol units consumed in the week prior to DNAm measurement. We began by evaluating two factors that could potentially affect the EpiScore’s prediction accuracy: (1) the training population and (2) the DNAm feature space.

To this end, we split the Generation Scotland into training (N = 8,684) and test sets (N = 8,033). Two choices for the training set were considered: everyone (N = 8,684) and the subset whose self-reported alcohol consumption in the previous week was reflective of a “normal week” (N = 5,618). The test set (N = 8,033) included all individuals, irrespective of the self-reported amount being more, less or about the same as normal. We trained predictors on either the full methylome (386,399 CpGs after limiting measured features to those also present in the Illumina 450K array for wider applicability) or 3,999 CpGs with previous evidence of an association to alcohol consumption in three recent EWASs that excluded Generation Scotland^9,15,29^ (see **Methods, Figure 1**). This resulted in a total of four EpiScores (**Figure 1**).

**Figure 1.**
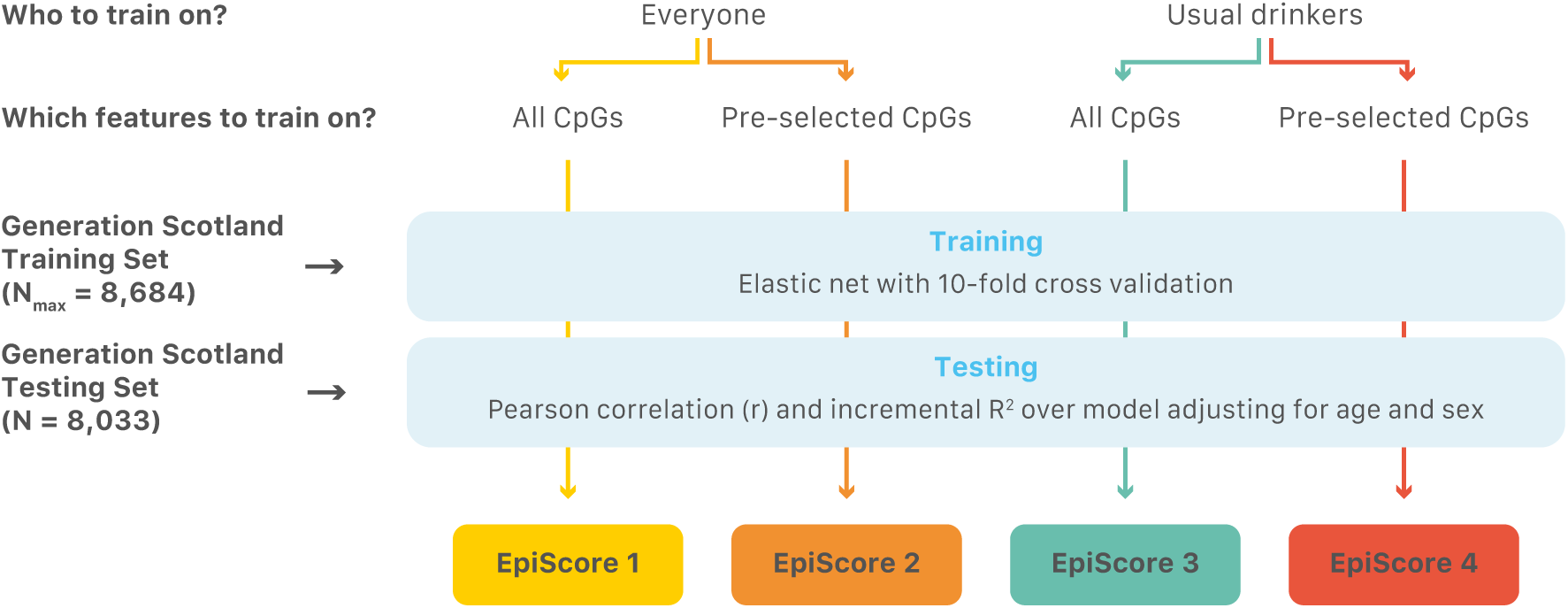
EpiScore predictor creation diagram.

EpiScore prediction performance was assessed by Pearson correlations (r) between self-reported alcohol consumption units per week and the EpiScore, as well as by calculating the incremental R^2^ upon the addition of the EpiScore to a linear regression model adjusting for age and sex, in the test set. We found that predictors trained on pre-filtered CpGs ahead of elastic net outperformed those trained on all CpGs, for both models trained on everyone as well as models trained on the subset whose consumption in the last week was noted to reflect most normal weeks (**Table 1, Supplementary Figure 4**). We further found that training on everyone as opposed to the subset whose drinking patterns in the previous 7 days reflected a normal week also increased prediction performance (**Table 1**, **Supplementary Figure 4**). This was found to likely be due to a larger sample size in training, as matching the training of the everyone subset to the same size as that of “normal week” drinkers (N = 5,618) returned very similar performance metrics between the two predictors (r = 0.43 and incremental R^2^ = 16.86, considering filtered CpGs in training).

**Table 1.** Predictive performance and number of features of four EpiScores generated using elastic net regression. Predictive performance was assessed in a holdout subset (test set) of Generation Scotland via Pearson correlations (r) and incremental R^2^ upon the addition of the EpiScore to a linear regression model for log alcohol units (+1) adjusting for age and sex. All EpiScores were marginally associated with self-reported alcohol consumption (P < 2.2 x 10^-16^). P-values taken for the EpiScore from the age- and sex-adjusted linear regression model.

| Training | CpGs | Elastic net N<br>selected<br>features | Everyone<br>(N = 8,033) |  | Normal<br>Week<br>(N = 4,888) |  | More than<br>normal<br>(N = 1,920) |  | Less than<br>normal<br>(N = 834) |  |
| --- | --- | --- | --- | --- | --- | --- | --- | --- | --- | --- |
| | | | $r$ | $R^2$ | $r$ | $R^2$ | $r$ | $R^2$ | $r$ | $R^2$ |
| Everyone<br>(N = 8,684) | All | 551 | 0.42 | 16.2 | 0.47 | 20.1 | 0.38 | 13.6 | 0.31 | 9.6 |
|  | Pre-selected | 343 | 0.44 | 18.2 | 0.50 | 22.6 | 0.40 | 15.4 | 0.31 | 9.6 |
| “Normal week”<br>Drinkers<br>(N = 5,618) | All | 460 | 0.40 | 15.3 | 0.45 | 19.0 | 0.36 | 13.4 | 0.30 | 9.2 |
|  | Pre-selected | 360 | 0.42 | 16.9 | 0.48 | 21.6 | 0.39 | 14.8 | 0.29 | 8.7 |

### Prediction performance by different drinking behaviours

If the methylome is only able to capture recent exposure to alcohol then our predictors should showcase differential performance if a person had deviated from their normal alcohol units consumed in a given week (drinking more or less than normal). We therefore evaluated the four EpiScores in the Generation Scotland test set, considering participants who reported their alcohol consumption was similar to a normal week versus those reporting having consumed more or less than normal over the past week (N = 7,642/8,033 – status not recorded for 391 individuals). This consisted of 4,888 “normal week” drinkers, 1,920 people who drank more than usual that week, and 834 people who drank less than usual. We found that the predictors performed best in the “normal week” drinkers, followed by those who had drank more or less than normal that week (**Table 1**, **Supplementary Figure 5**).

### EpiScore trained in all of Generation Scotland and tested in external cohorts

Having established that training on everyone and not just “normal week” drinkers in Generation Scotland, as well as pre-filtering features ahead of training optimised predictive performance, we trained our final model in this manner making use of the full cohort (N = 16,717). This returned an EpiScore consisting of 659 features (**Supplementary Table 3)**. Predictive performance was evaluated in the Lothian Birth Cohorts of 1921 and 1936 (N = 436 and 895 respectively, see **Methods**, **Supplementary Table 4, Table 2**). To further replicate our results, predictive performance was also tested in the external ALSPAC (5 cohort subsets, N_TOTAL_ = 4,083, ranging from 476 to 1,482 per cohort) and the Sister Study cohorts (2 cohort subsets, N_TOTAL_ = 5,119, with N = 2,770 and 2,349 per cohort respectively, see **Methods, Table 2**).

**Table 2.** EpiScore performance metrics in the Lothian Birth Cohorts, ALSPAC, and Sister Study cohorts. Model trained on all of Generation Scotland (N = 16,717). Cohort demographic metrics, as well as EpiScore performance metrics (r and incremental R^2^ over model adjusting for age and sex) are shown. LBC = Lothian Birth Cohort; SS = Sister Study; ALSPAC = Avon Longitudinal Study of Adults and Children; 15up = 15-17 year olds, measured either in 450K or EPIC arrays; F24 = 24 year olds; FOF: fathers in midlife; FOM: mothers in midlife; UPW: units per week.

|  | Cohort | N | Age (Mean, (SD)) | N Males (%) | Alcohol UPW (Median, [IQR]) | r | R<sup>2</sup> |
| --- | --- | --- | --- | --- | --- | --- | --- |
| LBC | LBC1921 | 436 | 79.1 (0.6) | 173 (39.7) | 1.0 [0.5, 7.0] | 0.41 | 17.9 |
|  | LBC1936 | 895 | 69.6 (0.8) | 453 (50.6) | 5.0 [0.5, 14.0] | 0.42 | 16.6 |
| ALSPAC | 15up 450K | 670 | 17.4 (0.9) | 311 (46.4) | 3.0 [1.6, 6.9] | 0.11 | 1.3 |
|  | 15up EPIC | 1,482 | 17.8 (0.4) | 704 (47.5) | 4.3 [1.3, 6.9] | 0.11 | 1.2 |
|  | F24 | 804 | 24.4 (0.8) | 409 (50.9) | 4.7 [2.3, 10.0] | 0.30 | 9.3 |
|  | FOF | 476 | 53.3 (5.3) | 476 (100.0) | 6.9 [3.0, 10.0] | 0.45 | 20 |
|  | FOM | 651 | 47.4 (4.4) | 0 (0.0) | 4.3 [1.3, 6.9] | 0.33 | 10.7 |
| SS | 450K | 2,770 | 57.0 (8.8) | 0 (0.0) | 1.0 [0.0, 4.0] | 0.53 | 28.1 |
|  | EPIC | 2,349 | 55.5 (8.7) | 0 (0.0) | 0.5 [0.0, 3.0] | 0.51 | 26.9 |
|  | EPIC nH White | 1,553 | 56.4 (8.8) | 0 (0.0) | 0.9 [0.1, 4.0] | 0.51 | 26.6 |
|  | EPIC Black | 796 | 53.7 (8.3) | 0 (0.0) | 0.1 [0.0, 1.0] | 0.37 | 14.4 |

Our EpiScore correlated with self-reported alcohol consumption (r_LBC21_ = 0.41, r_LBC36_ = 0.42), and the marginal association remained after adjusting by age and sex (P_EpiScore_ < 2.2 x 10^-16^ in both cohorts. The EpiScore had an incremental R^2^ (over a linear regression model adjusting for age and sex) of 17.9% in LBC1921 and of 16.6% in LBC1936 (**Table 2, Supplementary Figure 6**). This outperforms a previously published alcohol consumption EpiScore trained on a subset of N = 2,819 in Generation Scotland^21^, which presented an incremental R^2^ over a model adjusting for age and sex of 6.3% in LBC1921 and 10.6% in LBC1936.

Considering the five ALSPAC cohort timepoints (15up 450/EPIC: 15-17 year olds measured on either 450K or EPIC Illumina chips, F24: 24 year olds, FOM: mothers in midlife, and FOF: fathers in midlife), the EpiScore correlation with self-reported alcohol consumption ranged from r = 0.11 to 0.45, with an incremental R^2^ over a linear regression model adjusting for age and sex ranging from 1.2% to 20%. Notably, the worst performing subsets were made up of young individuals (mean age less <18). The two Sister Study cohort subsets (one measured with 450K array and another with EPIC array) showed correlations of r = 0.53 and 0.51, and with an incremental R^2^ = 28.1% and 26.9%, respectively (**Table 2**, **Supplementary Figure 7 and 8**). The mean age in the Sister Study cohorts was approximately 56 years old.

### EpiScore categorization of heavy drinkers

Next, we evaluated our EpiScore’s ability to discriminate heavy drinkers versus light-to-moderate drinkers in LBC1921 and 1936. We dichotomized the alcohol consumption phenotype into ‘cases’ (heavy drinkers) if they had drunk over 21 or 14 units in the measured week for males and females, respectively, and as ‘controls’ if otherwise^30^. A total of 33 (7.6% out of N = 436) and 150 (16.8% out of N = 895) heavy drinkers were present in LBC1921 and LBC1936, respectively (**Supplementary Table 3**).

We calculated the area under the receiver operating characteristic curve (AUROC) for the binary drinking phenotype (see **Methods**) and found good-to-excellent discrimination of heavy drinkers versus light-to-moderate drinkers, with AUROC 0.9 (CI 95% 0.85, 0.95) in LBC1921 and AUROC 0.78 (CI 95% 0.74, 0.82) in LBC1936 (**Figure 2**, **Table 2**). This surpasses the classification performance of a previously published predictor trained on a subset of N = 5,087 in Generation Scotland^21^, which returned an AUROC = 0.77 (CI 95% 0.68, 0.86) and AUROC = 0.73 (CI 95% 0.68, 0.77) in LBC1921 and LBC1936, respectively (**Table 2**).

**Figure 2.**
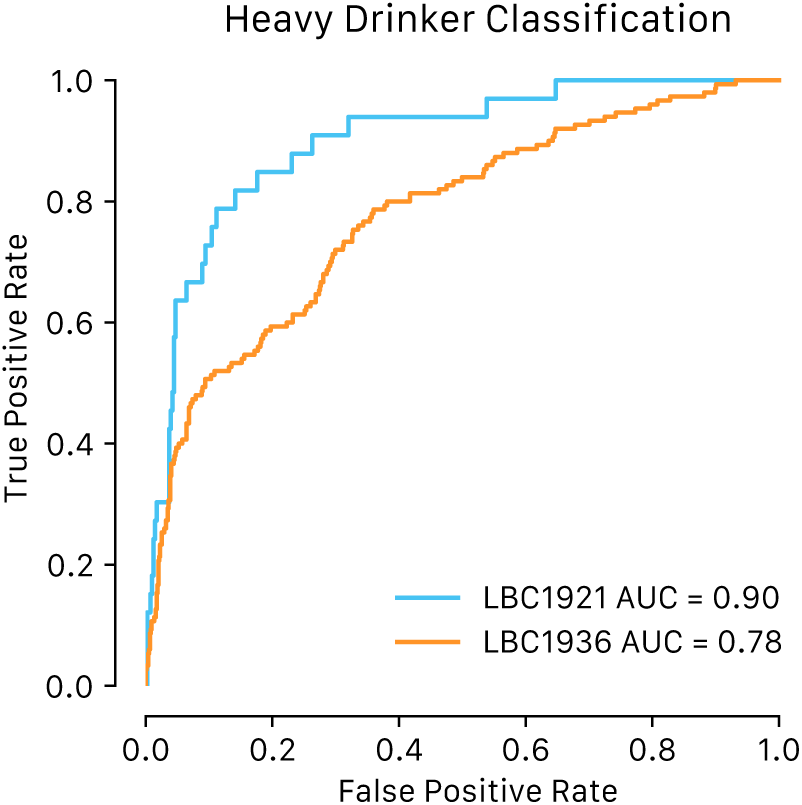
AUC analysis of alcohol consumption prediction in the Lothian Birth Cohorts of 1921 and 1936. Shown are area under the receiver operating characteristic curves for classifying dichotomized alcohol consumption (heavy drinkers - >14 units per week for females or >21 units per week for males - versus non-or light-moderate drinkers).

Given the unbalanced nature of this phenotype, which can lead to an over-optimistic AUROC, we also calculated the area under the precision recall curve (AUPRC), with AUPRC = 0.44 in LBC1921 and AUPRC = 0.47 in LBC1936 (**Supplementary Figure 9**).

### Measured alcohol consumption and EpiScore associations

Given that alcohol consumption is not an easy phenotype to quantify reliably via self-report, an EpiScore may act as an alternative or complementary measure in the search for associations with disease and other factors. We therefore tested for associations between self-reported alcohol consumption or our alcohol EpiScore and a number of lifestyle/health/socioeconomic factors (smoking, years of education, walking speed, grip strength, occupational social class), biomarkers (cholesterol, triglycerides), self-reported disease history (cardiovascular, cerebrovascular, neoplasia, hypertension, diabetes, thyroid, depression, anxiety), MRI brain variables, and survival in the Lothian Birth Cohorts using a series of linear and logistic regression models, adjusting for age and sex (see **Methods, Supplementary Table 5**).

Although not always statistically significant (FDR corrected P < 0.05), the EpiScore and self-reported alcohol consumption presented consistent positive associations across both LBC cohorts with number of packs smoked per day and smoking status, years in education, and cholesterol levels. On the other hand, both measures were consistently negatively associated with occupational social class, diabetes prevalence, and anxiety scores. Further, both presented HR > 1 in relation to time to all-cause mortality.

A small number of the explored associations were statistically significant (FDR corrected P < 0.05). Considering lifestyle and cognitive traits, the EpiScore was associated with smoking status in LBC1936 (standardised β = 0.117, P_FDR_ = 0.009) and with occupational social class in LBC1921 (β = −0.105, FDR P = 0.024). On the other hand, a higher self-reported alcohol consumption was associated with smoking status in LBC1921 (β = 0.17, P_FDR_ = 0.011). Considering disease history, the EpiScore was positively associated with high blood pressure in LBC1936 (OR_per SD of the EpiScore_ = 1.22, P_FDR_ = 0.012). Self-reported alcohol consumption was not significantly associated with any of the disease histories considered here. Further, we found a significant association between our EpiScore and time to all-cause mortality in LBC1936 (HR = 1.16 [95% CI 1.05, 1.28]).

All brain MRI variables tested here were found to be significantly associated with the EpiScore in LBC1936, and were not found to be significantly associated with self-reported alcohol consumption (**Figure 3**). These included negative associations with total brain volume (β = −0.044, P_FDR_ = 0.012), grey matter volume (β = −0.074, P_FDR_ = 0.001), and normal appearing white matter volume (β = −0.064, P_FDR_ = 0.012), and a positive association with white matter hyperintensity volume (β = 0.120, P_FDR_ = 0.012).

**Figure 3.**
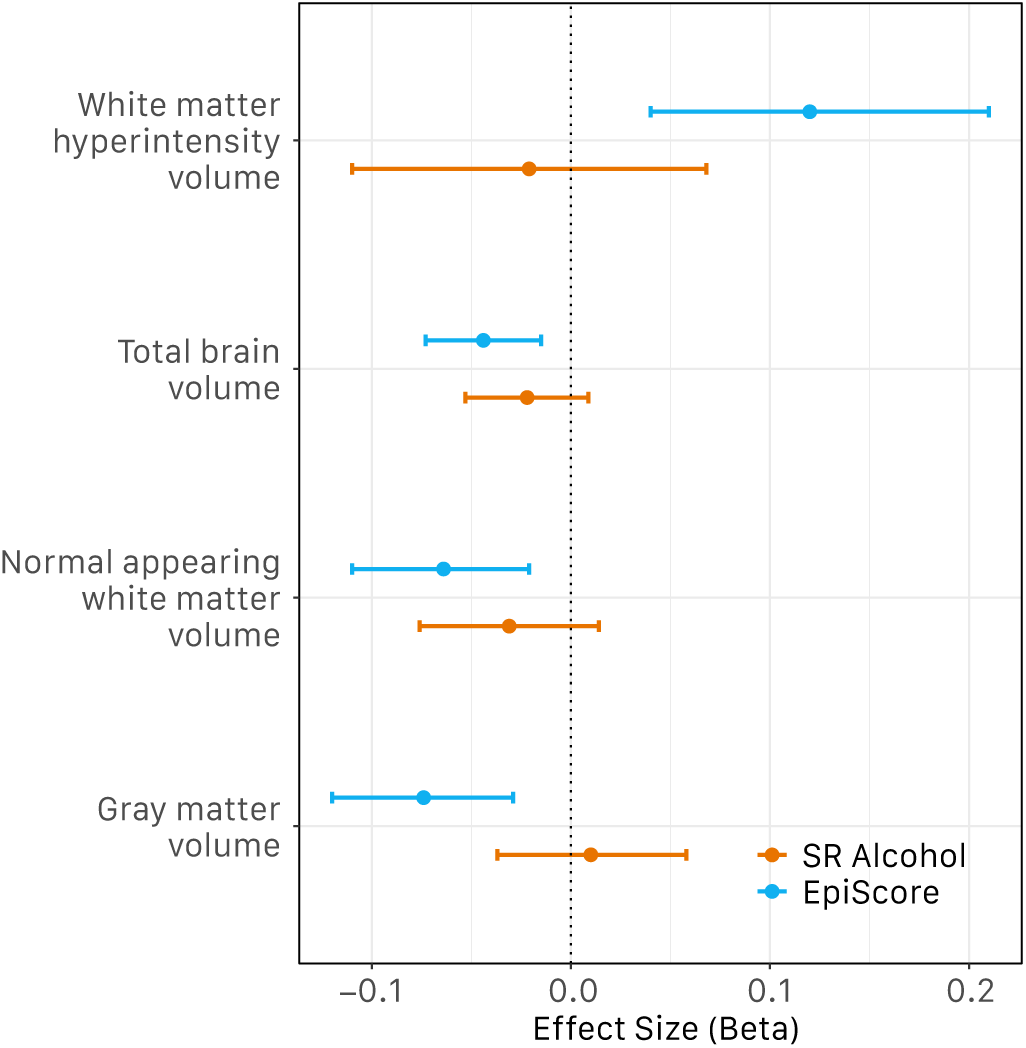
Self-reported (SR) alcohol consumption and alcohol EpiScore associations with global brain imaging in LBC1936. Standardized effect sizes from age and sex adjusted linear regression models shown along with 95% confidence intervals.

### Sex-specific EpiScore performance

Given the differences in alcohol consumption between males and females, we explored sex-specific models. We trained EpiScores in Generation Scotland, considering the pre-selected features described previously, as well as matching sample sizes (N = 6,958, given this is the size of the smallest subset: N_males_), in a sex-agnostic manner (equal number of males and females, N = 3,479 each), in a female-specific model, and in a male-specific model (see **Methods**). We then assessed the prediction performance of the EpiScores in the Lothian Birth Cohorts in three different ways: (1) applying the sex-agnostic EpiScore to everyone (sex-agnostic prediction), (2) applying the female-specific EpiScore to female samples and the male-specific predictor to male samples (same-sex prediction), and (3) applying the female-specific EpiScore to male samples, and the male-specific EpiScore to female samples (opposite-sex prediction).

We found that prediction performance, as measured by r and incremental R^2^ over a model accounting for age and sex, did not vary greatly across EpiScores (**Supplementary Figure 10**, **Table 3**) in both LBC1921 and LBC1936.

**Table 3.** Sex-specific EpiScore performance metrics in the Lothian Birth Cohorts. Model trained in Generation Scotland, matching sex sample size (sex-agnostic, same-sex, and opposite-sex, N = 6,958). EpiScore performance metrics (r and incremental R^2^ over model adjusting for age and sex) are shown for LBC1921 and LBC1936. Also shown is classification performance for heavy drinkers (AUC for ROC curve, and its corresponding 95% confidence interval). All EpiScores associated with measured alcohol consumption with P < 2.2 x 10^-16^. Sex-agnostic: Sex-Agn. Opposite-sex: Oppo-sex.

| Model | Elastic net<br>N selected features | LBC1921 |  |  | LBC1936 |  |  |
| --- | --- | --- | --- | --- | --- | --- | --- |
|  |  | r | R<sup>2</sup> | AUC [95% CI] | r | R<sup>2</sup> | AUC [95% CI] |
| **Sex-Agn** | 423 | 0.36 | 20.6 | 0.88 [0.81-0.94] | 0.39 | 14.4 | 0.76 [0.72-0.81] |
| **Same-sex** | 445 | 0.37 | 21.0 | 0.87 [0.81-0.93] | 0.42 | 15.1 | 0.78 [0.73-0.82] |
| **Oppo-sex** | 370 | 0.38 | 21.2 | 0.87 [0.8-0.94] | 0.41 | 14.5 | 0.76 [0.72-0.8] |

We assessed the performance of these EpiScores to classify heavy drinkers, as described before. No large performance differences were found between them (**Table 3**).

### Alcohol consumption variance explained by the methylome and EWAS

Next, we determined the proportion of variance in the alcohol consumption phenotype that can be explained by all CpG sites measured on a DNAm array (more specifically, the Illumina EPIC array, consisting of 752,722 CpGs after QC). To do this, we fitted a Bayesian sparse regression model and performed a variance partitioning analysis using BayesR+ (see **Methods**). BayesR+ has been shown to implicitly control for white cell proportions, which are typically estimated from the DNAm data, related participants, and other unknown confounders^31^. Three mixture distributions were specified, corresponding to possible small, medium and large effect sizes for the CpGs (explaining 0.01%, 0.1% and 1% of the variance, respectively). We fit models using (i) “normal pattern” drinkers in Generation Scotland, and (ii) the full Generation Scotland cohort. Our analyses found that 45.0% (95% Credible Interval 39.7%, 50.5%) and 49.3% (95% Credible Interval 44.3%, 54.4%) of alcohol consumption was explained by all CpGs in models with “normal week” drinkers (those whose self-reported alcohol consumption was consistent with their normal drinking behaviour) and the full cohort, respectively.

In addition to the variance components analysis, BayesR+ simultaneously conducts an epigenome-wide association study (EWAS - see **Methods**). This assesses the association between each CpG and the outcome. We found a total of four and six lead CpGs with a posterior inclusion probability (PIP) greater than 0.95 (**Figure 3**, **Supplementary Table 6**) in models considering just “normal week” drinkers and the full cohort, respectively. Two CpGs had a PIP greater than 0.95 in both models, and a total of eight unique lead CpGs were found.

**Figure 3.**
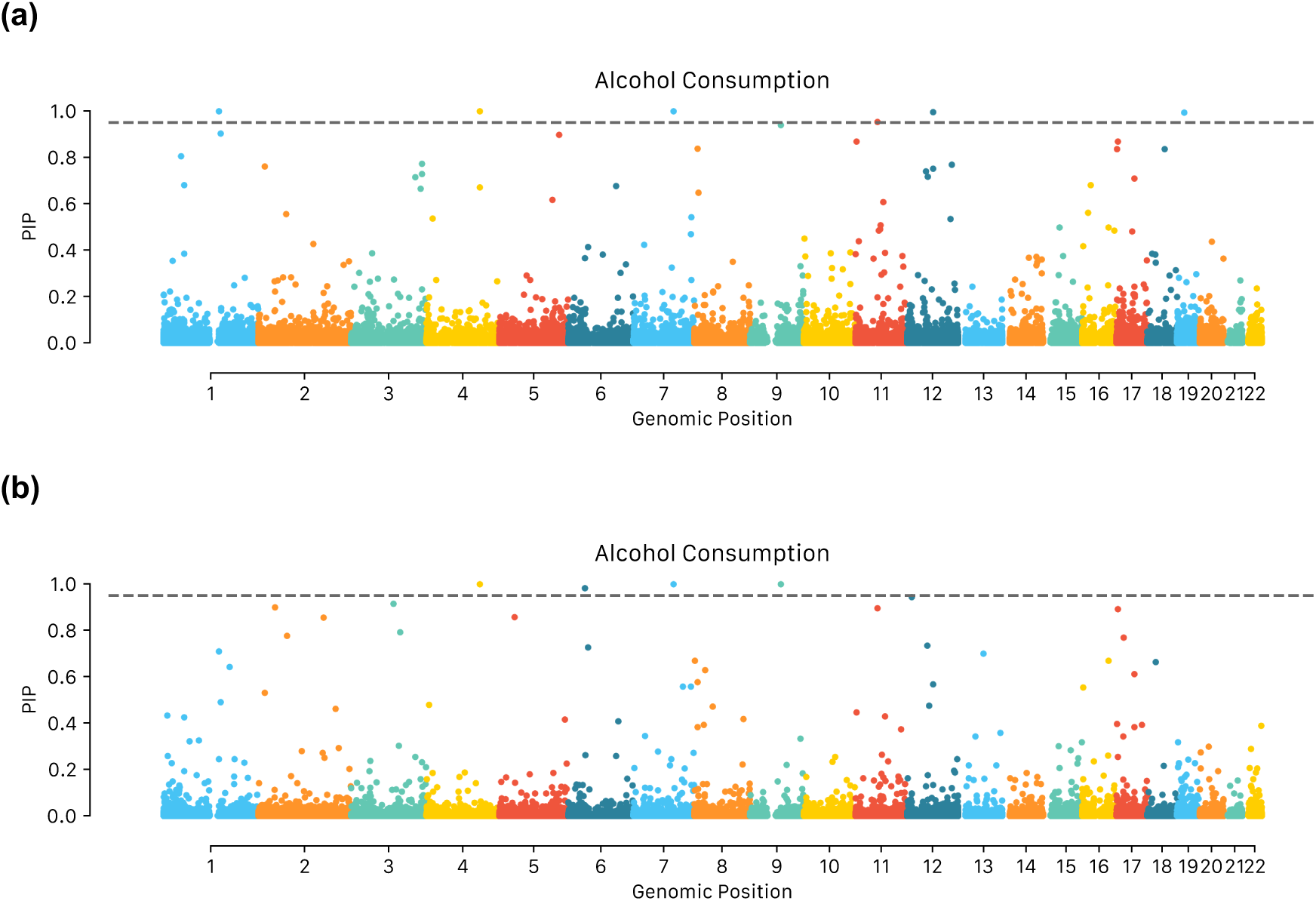
EWAS of alcohol consumption Manhattan plot. Model using 1) usual drinkers and 2) full cohort. Threshold line set at posterior inclusion probability (PIP) = 0.95.

We queried the EWAS catalog (accessed 2^nd^ July, 2023) for the eight aforementioned CpG sites and found that three had been previously linked to alcohol consumption, while one had been previously linked to alcohol withdrawal recovery (**Supplementary Table 7**). This search is not exhaustive, as not all studies deposit data in this resource. Indeed, six of the eight CpG sites were found in the largest previously published alcohol consumption EWAS making use of Generation Scotland data^11^ (all but cg03741185 and cg06053623). Seven of the eight CpGs were found to be associated with at least one other trait in the EWAS catalog, including: age, prevalent type 2 diabetes, serum high-density cholesterol, gestational age, serum triglycerides, blood pressure, BMI, and others.

Our lead CpGs mapped to the genes *POLR3GL, SLC7A11, SRPK2, PSAT1, IL12RB1, SLC43A1,* and *LOC100132354*. *IL12RB1* has not been previously linked to alcohol consumption, based on EWAS Catalog output and the previously largest GS-based alcohol consumption EWAS.

## Discussion

Excessive alcohol consumption is one of the most important contributors to the global burden of disease, with important associations to conditions including cardiovascular disease, cancer, and more^3–5^. Alcohol has further been associated with DNAm differences via multiple mechanisms^32^ and as such, the altered methylome could offer clues into alcohol-disease links.

Here, we report a new EpiScore which improves upon a previous epigenetic predictor of alcohol consumption trained in Generation Scotland, finding a correlation between our predictor and measured alcohol of up to 0.47, and an incremental R^2^ over a model considering just age and sex of up to 22.3% in the Lothian Birth Cohorts, compared to an incremental R^2^ of 12.5% obtained with our previously published predictor^21^. We further replicated our findings in the ALSPAC and Sister Study cohorts, where we found a correlation with measured alcohol of up to 0.53, and an incremental R^2^ of up to 28.1%. These tests highlighted that the EpiScore performs considerably better in middle-aged to older individuals than in individuals aged under 20. In addition, we found that this EpiScore classified heavy drinkers within the Lothian Birth Cohorts with an AUC of up to 0.9 (CI 95% 0.85, 0.95), compared to an AUC of 0.77 (CI 95% 0.68, 86) with the previously published predictor^21^. This EpiScore could offer an alternative and potentially more objective measure to self-reported alcohol consumption for downstream analyses with health outcomes.

Our alcohol EpiScore also associated negatively with white matter, grey matter, and total brain volume, and positively with white matter hyperintensity volume, whereas self-reported alcohol consumption did not. Chronic alcohol use is associated with changes in brain structure and connectivity^33^, and previous studies have reported links between higher alcohol consumption and lower white and grey matter volume^34^, as well as with higher white matter hyperintensity volume^35^. A recent study making use of the UK Biobank brain MRI data (N = 36,585)^2^ found that self-reported alcohol consumption was associated negatively and slightly non-linearly with both white matter and grey matter volumes, after accounting for covariates including age, sex, and BMI. The authors report that consuming as few as 1-2 alcoholic drinks daily was associated with decreased brain volume. The lack of significant associations between self-reported alcohol consumption and brain volumes in our study could be due to lack of statistical power, or a difference in accuracy between the measurements used in the two studies.

We found that training on everyone yielded a better predictor than training just on those whose self-reported drinking in the week prior to the blood draw and questionnaire was similar to that during a normal week. This could be due to larger sample sizes with this hypothesis being supported by similar performance metrics when training on equal sample sizes for both everyone (more than normal, less than normal, and “normal week”) and “normal week” drinkers subsets. Further, we found that our predictor performed best when testing on “normal week” drinkers, as opposed to samples that reported abnormal drinking (more or less than normal) in the week consumption was measured. This suggests that the alcohol methylome is dynamic and reversible. Indeed, a recent study found that a large number of CpG sites that were found to be associated with alcohol consumption presented differential methylation between former and current drinkers, and found that alcohol-related hypomethylation is largely reversible upon cessation^15^. Our results suggest that changes to the methylome could be observed in short time-frames, but longitudinal data with frequent time points would be needed to confirm this.

Previous studies have found that pre-filtering ahead of elastic net greatly improves predictor performance when using this training method^36^. Our current results echo this, with a significant increase in prediction accuracy found when training on CpGs with a previously established association to alcohol consumption (**Table 1**). This could be due to less overfitting in the training set alongside the screening out of CpGs with low intra-sample variability due to technical variance^37,38^.

Alcohol consumption patterns and alcohol-related complications differ between the sexes^34,39,40^. In addition, sex differences in the methylome have been described^41^. Here we found that sex-specific EpiScores yielded very similar results when matching sample sizes and comparing to sex-agnostic models. This could suggest that (1) no major differences exist, (2) an EpiScore does not reflect alcohol-related biological differences between the sexes, or (3) insufficient sample size was used in training to detect smaller sex-specific patterns. We also show the importance of sample size when training EpiScores; compared to a previous EpiScore, trained in 2,819 unrelated Generation Scotland volunteers with “normal week” drinking patterns, our new score explained 1.7- and 2.8-fold more variance in a self-reported alcohol consumption phenotype from the Lothian Birth Cohorts of 1936 and 1921, respectively.

To gain insights into the associations between individual CpG loci and alcohol, we performed the largest single-cohort EWAS of alcohol consumption to date. We found 8 sentinel loci, which mapped to 7 unique genes. Three of the genes the CpGs map to, which have already been reported to be associated to alcohol consumption in previous EWAS efforts^9,11,15,42^, are part of the aminotransferase family (*SLC7A11, PSAT1,* and *SLC43A1*). Alcohol is known to disrupt protein metabolism and amino acid transport^43,44^ and *SLC7A11*’s role in the liver– brain-axis in alcohol-related disease and potential as a future drug target has been described^11^. One of our strongest CpG associations (cg26774981) mapped to the *SRPK2* gene, a kinase that controls alternative splicing. A recent paper found the regulation of alternative splicing by SRPK2 is implicated in lipogenesis in humans with alcohol-associated liver disease, thus making it a potential drug target^45^. One of the seven genes mapping to the CpG loci we identified has not been linked to alcohol consumption: a type I transmembrane protein of the hemopoietin receptor superfamily (*IL12RB1*). Future work is needed to replicate these findings, and to understand their potential role in alcohol-mediated disease etiology.

Our study has several limitations. Firstly, the majority of the Generation Scotland and Lothian Birth Cohorts are of White British ancestry, which could lead to biases and difficulty translating these results to other population. However, the EpiScore performed as well or better in two external cohorts of diverse age ranges (ALSPAC) and ancestries (Sister Study). Secondly, as has been discussed previously, this study is based on an imperfect phenotype. Indeed, self-reporting has its limitations, and further details regarding alcohol consumption, such as a breakdown of type of drink consumed (beer, wine, spirits), could help further untangle its relationship with human health and the methylome. Thirdly, DNAm was measured in whole blood, and therefore these results may not apply to all blood cell types or other mechanistically relevant tissues such as brain. Fourth, whilst our EpiScore led to good discrimination between heavy and non-heavy drinkers, the separation is not perfect, and prediction could benefit from the use of more complex algorithms (e.g. to capture non-linear associations between DNAm and alcohol consumption) as well as larger sample sizes.

Overall, our study expands on the existing literature on alcohol consumption and its relation to the DNA methylation, including the creation of an improved epigenetic predictor as well as new insights into links between alcohol and the methylome.

## Methods

### The Generation Scotland Cohort

#### DNA methylation

DNA methylation in blood at baseline (recruitment) was quantified for 18,413 Generation Scotland participants across three separate sets (N_Set1_ = 5,087, N_Set2_ = 4,450, N_Set3_ = 8,876) using the Illumina MethylationEPIC (850K) array. Individuals in Set 1 included a mixture of related and unrelated individuals. Set 2 comprised individuals unrelated to each other and to those in Set 1. Set 3 contained a mix of related individuals – both to each other and to those in Sets 1 and 2 – and included all remaining samples available for analysis. Methylation data was processed across 121 experimental batches (N_Batches. Set1_ = 31, N_Batches, Set2_ = 30, N_Batches, Set3_ = 60).

Quality control details have been reported previously^46,47^. Briefly, probes were removed based on (i) outliers from visual inspection of the log median intensity of the methylated versus unmethylated signal per array, (ii) a bead count < 3 in more than 5% of samples, (iii) ≥ 5% of samples having a detection *p*-value > 0.05, (iv) if they pertained to the sex chromosomes, (v) if they overlapped with SNPs, and/or (vi) if present in potential cross-hybridizing locations^48^. Samples were removed (i) if there was a mismatch between their predicted sex and recorded sex, (ii) if ≥ 1% of CpGs had a detection *p*-value > 0.05, (iii) if sample was not blood-based, and/or (iv) if participant responded “yes” to all self-reported diseases in questionnaires. A total of 752,722 CpGs remained after QC. Missing values were imputed using the mean of each CpG across all samples. Dasen normalisation^49^ was performed across all individuals.

#### Alcohol consumption data

Self-reported alcohol consumption was measured at baseline via questionnaires to obtain the number of units consumed in previous week (unit definition as per UK National Health Service: 8g/10ml of pure alcohol). Participants were also asked whether this was their usual drinking amount, or whether they had consumed more or less than normal. A total of 16,717 individuals had non-missing alcohol consumption data and methylation data – the rest of participants were excluded from this study (after exclusion, N_Set1_ = 4,576, N_Set2_ = 4,108, and N_Set3_ = 8,033 individuals were left in sets 1, 2 and 3, respectively). Of these, 10,506 marked this quantity as representative of their typical weekly consumption, and 3,756 stated this quantity was more than normal and 1,622 less than normal (**Supplementary Table 1, Supplementary Figure 1**).

### Lothian Birth Cohorts

#### Overview

LBC1921 and LBC1936 are longitudinal studies of ageing on individuals born in 1921 and 1936, respectively^23^. Study participants completed the Scottish Mental Surveys of 1932 and 1947 at approximately age 11 years old and were living in the Lothian area of Scotland at the time of recruitment in later life.

#### DNA methylation

Blood samples considered here were collected at around age 79 for LBC1921, and at around age 70 for LBC1936. DNA methylation was quantified using the Illumina HumanMethylation450K array, for a total of 692 (up to 3 repeated measurements from 469 individuals) and 2,796 (up to 4 repeated measurements from 1,043 individuals) samples from LBC1921 and LBC1936 respectively. Quality control details have been reported previously^50^. Briefly, probes were removed (i) if they presented a low (< 95%) detection rate with *p-*value < 0.01, and/or (ii) if they presented inadequate hybridization, bisulfite conversion, nucleotide extension, or staining signal, as assessed by manual inspection. Samples were removed (i) if they presented a low call rate (<450,000 probes detected at *p*-value < 0.01) and/or (ii) if predicted sex did not match reported sex. Missing values were replaced with 0.

#### Self-reported alcohol consumption

Participants were asked about their usual alcohol consumption, including number of times alcohol is consumed per week, normal alcohol consumption, typical drink of choice, and glasses/pints consumed on average. From this information, alcohol consumption in units consumed per week was derived. A total of 436 and 895 individuals had non-missing alcohol consumption and methylome data available in LBC1921 and LBC1936 baseline, respectively, and were considered in this study.

### ALSPAC

#### Overview

The Avon Longitudinal Study of Parents and Children (ALSPAC) is a cohort study of pregnant women resident in Avon, UK with expected dates of delivery between 1st April 1991 and 31^st^ December 1992^25,26^. Among these, 20,248 pregnancies were identified as being eligible and the initial number of pregnancies enrolled was 14,541 resulting in 14,062 live births and 13,988 children who were alive at 1 year of age. At the start of the study, mothers invited their partners to complete questionnaires. In total, 12,1113 partners have provided data and 3807 are currently formally enrolled. As part of Accessible Resource for Integrated Epigenomic Studies (ARIES)^51,52^, a sub-sample ALSPAC children, mothers and partners had DNAm assayed using the Illumina Infinium HumanMethylation450 or MethylationEPIC Beadchip array from peripheral blood samples collected at multiple time points from birth to middle age. The present study used DNAm measured from peripheral blood samples collected from ALSPAC children at ages 15-17 (time-point ‘15up’) and 24 (time-point ‘F24’)^53^, and from ALSPAC mothers and partners^54^ 18 years after the study pregnancy. Study data were collected and managed using REDCap electronic data capture tools hosted at the University of Bristol. REDCap (Research Electronic Data Capture) is a secure, web-based software platform designed to support data capture for research studies^55^. Please note that the study website contains details of all the data that is available through a fully searchable

#### DNA methylation

Illumina Infinium HumanMethylation450 and MethylationEPIC Beadchip arrays were used to assess genome-wide DNAm patterns in peripheral blood. Samples across different time-points were distributed in a semi-random manner across slides in order to mitigate batch effects. Data pre-processing and normalization was performed using the R package meffil as previously described^52^. Samples with large numbers of undetected probe signals ( were removed, along with those that had sex or genotype mismatches. Probes undetected in more than 20% of samples were excluded.

#### Self-reported alcohol consumption

Alcohol consumption was measured as the estimated number of units consumed on average during the week the year before blood sample collection for DNAm analysis. Consumption was estimated multiplying alcohol intake frequency per week by intake quantity. Frequency was assessed by the question ‘How often do you have a drink containing alcohol’, with possible responses including ‘Never’, ‘Monthly or less’, ‘2 to 4 times a month’, ‘2 to 4 times a week’ and ‘4 or more times a week’. ‘Never’ drinking respondents were considered non-drinkers and were included in all primary analyses. Quantity was assessed by asking the number of drinks consumed where ‘one drink referred to ½pint of beer/cider, a small (125 ml) glass of wine or a single (25 ml) measure of spirit’, each of which is roughly equivalent to one UK alcohol unit (8g of ethanol).

### Sister Study

#### Overview

The Sister Study is a US-nationwide prospective cohort study of 50,884 women enrolled between 2003 and 2009; women were eligible for enrollment if they resided in the United States and were breast cancer-free themselves but had a biological sister who was previously diagnosed. As part of study enrollment when all women were breast cancer-free, women completed self-reported questionnaires and an in-home visit where a whole blood sample was collected. Information about obtaining data from the Sister Study can be found at: https://sisterstudy.niehs.nih.gov/English/coll-data.htm.

#### DNA methylation

Two case-cohort samples of women were selected for DNAm profiling. In 2014, blood DNA samples from 2,878 self-identified non-Hispanic White women were assayed on the Infinium HumanMethylation450 BeadChip^56^. This sample included 1,542 women who were diagnosed with breast cancer in the years following enrollment (mean time to diagnosis: 4 years). In 2019, blood DNA samples from 2,599 self-identified Black (Hispanic and non-Hispanic) and non-Hispanic White women were assayed on the Infinium MethylationEPIC v1 BeadChip^57^. This sample included 999 women who were diagnosed with breast cancer in the years following enrollment (mean time to diagnosis: 5 years). Self-identified Hispanic and non-Hispanic Black women were over-sampled for DNAm profiling in order to maximize the racial and ethnic diversity of the MethylationEPIC sample.

For both DNAm samples, DNAm data were preprocessed using the *ENmix* software pipeline, which included background correction, dye-bias correction, inter-array normalization, and probe-type bias correlation^58–60^. Samples were excluded if they did not meet quality control measure including bisulfate intensity < 4,000, had greater than 5% of probes with low quality methylation values (detection P > 0.000001, < 3 beads, or values outside 3 times the interquartile range), or were outliers for their methylation beta value distributions. In total, 178 participants from the HumanMethylation450 sample and 250 participants from the MethylationEPIC sample were excluded for not meeting quality control measures.

#### Alcohol consumption

Participants’ history of alcohol consumption was obtained within 1 year of blood draw as part of a baseline questionnaire for alcohol use. Women reported information including the age at which they started and stopped drinking alcohol. The frequency of alcohol consumption was reported as days per week, month, or year by decade of life. The alcohol use variable used in this study was a derived variable that represented the average number of drinks per week over the last twelve months.

### EpiScore of alcohol consumption: who to train on, and how?

In an effort to assess the optimal cohort sample and feature space to train on, multiple EpiScores were assessed. The Generation Scotland cohort was divided into a training (sets 1 and 2, N = 8,684) and a testing dataset (set 3, N = 8,033). EpiScores were trained on the full training dataset, as well as just on the “normal week” drinkers (N = 5,618). Further, EpiScores were trained on the full methylome (386,399 CpGs after limiting measured features to those also present in the Illumina 450K array for wider applicability) or on a subset of 3,999 epigenome-wide significant CpGs (P < 3.6 x 10^-8^) that have been previously linked to alcohol consumption in three separate studies not using Generation Scotland^9,15,29^.

Elastic net penalized regression was used to train our EpiScores on log-transformed alcohol consumption + 1 (*glmnet* package in R, v4.1). CpG beta values in the training set were scaled to mean zero and unit variance ahead of elastic net. The L_1_, L_2_ mixing parameter was set at α = 0.5, and 10-fold cross validation was performed to select the shrinkage parameter (λ) that minimised the mean cross-validated prediction error.

Predictive performance for each EpiScore was assessed by projecting the latter into the testing dataset by multiplying each CpG by its estimated weight and performing summation, scaling CpG beta values beforehand to mean zero and unit variance. Pearson correlation (r) of the EpiScore with measured log alcohol consumption + 1, as well as the incremental R^2^ upon the addition of the EpiScore to a linear regression model adjusting for age and sex, were then calculated. EpiScore statistical significance was assessed considering the marginal test for the beta in the linear regression model adjusting for age and sex (t-test assessing whether beta is significantly different from zero).

### Training the EpiScore in Generation Scotland and testing in the Lothian Birth Cohorts, ALSPAC and Sister Study

Having established that training on all individuals with self-reported alcohol consumption data (regardless of whether this pattern reflected a typical week or was more or less than normal), and on a pre-filtered set of CpGs, yields the better performing EpiScore, we next trained on the full Generation Scotland cohort (N = 16,717). As with the creation of previous EpiScores, elastic net penalized regression was used with α = 0.5 and 10-fold CV. This EpiScore was then projected and tested on the Lothian Birth Cohorts of 1921 and 1936, ALSPAC, and the Sister Study. Its performance was again assessed via a Pearson correlation with self-reported alcohol consumption and the incremental R^2^ upon the addition of the EpiScore to a linear regression model adjusting for age and sex.

#### Categorization of heavy drinkers

To assess the heavy drinker classification performance of each EpiScore in the Lothian Birth Cohorts, self-reported alcohol consumption in the testing dataset was binarized (heavy drinker consuming over 14 or 21 units week for females and males, respectively – as per the health guidelines at the time of data collection^30^). Receiver Operating Characteristic (ROC) curves were then obtained for heavy/not-heavy alcohol consumption, and Areas Under the ROC (AUROC) curves and their corresponding 95% confidence intervals were estimated using the *pROC* package in R (v1.18.2). Precision Recall (PR) curves were also obtained and AUCs calculated using the *PRROC* package in R (1.3.1).

#### Sex-specific EpiScores

Sex-specific EpiScores were trained after matching sample sizes (thus ensuring larger sample sizes weren’t driving better prediction). Given that the smallest sex-stratified sample size was N = 6,958 (males), we trained male-specific EpiScore on the full male sample set, a female-specific EpiScore trained on a random subsample of 6,968 female participants, and a sex-agnostic EpiScore trained on equal numbers of males and females with overall sample size also being 6,958 (N_F_ = 3,479, N_M_ = 3,479).

To assess performance, using the same metrics and testing LBC dataset described previously, we tested the resulting EpiScores in three different ways: (1) a sex-specific manner by which predictions are obtained using each testing sample’s sex-specific EpiScore, (2) an opposite-sex manner, by which the EpiScore trained on the opposite sex of the testing sample is used to obtain predictions, and (3) a sex-agnostic manner, by which all samples, regardless of sex, are predicted using the EpiScore trained on both males and females.

### EpiScore and self-reported alcohol consumption associations in the Lothian Birth Cohorts

Associations between multiple phenotypes and self-reported alcohol consumption, as well as with our generated EpiScore trained on the full Generation Scotland cohort, were evaluated separately in LBC1921 and LBC1936. For each phenotype, linear regression models were run, adjusting for age, sex, and either self-reported alcohol consumption or the epigenetic predictor. Phenotypes considered included body mass index (BMI in kg/m^2^), hand grip strength (maximum of left and right hand measurements), self-reported years of education, self-reported smoking status (never smoker, ex-smoker, and current smoker), number of smoked packs per day, measured time taken to walk 6 meters (in seconds), occupation-based social class (measured as social grades based on highest reached occupation^61^), and depression and anxiety scores (HADS-D and HADS-A total from the Hospital Anxiety and Depression questionnaire^62^). Associations with blood biomarkers cholesterol and triglycerides were also assessed.

Self-reported alcohol and alcohol EpiScore associations with self-reported prevalent disease were evaluated using logistic regression, adjusting for age and sex. These included CVD, stroke, neoplasm, high blood pressure, diabetes, and thyroid dysfunction. Associations with time to all-cause mortality were assessed using a Cox proportional hazards model with age and sex as covariates, using the *survival* R package (v3.5), with time to all-cause mortality or censoring as the survival outcome.

Finally, associations with multiple brain imaging phenotypes measured in LBC1936 were considered. Briefly, Structural and diffusion tensor (DTI) MRI acquisition and processing in LBC1936 were performed at Wave 2 (age 73 years) according to an open-access protocol^63^. Total brain, grey matter and normal-appearing white matter (NAWM) volumes were calculated using a semi-automated multi-spectral fusion method^64^. Intracranial volume was determined semi-automatically using Analyze 11.0^TM^. Total brain, grey matter, and white matter volume measurements were scaled to mean zero and unit variance, and associations with self-reported alcohol consumption and the alcohol EpiScore were assessed via linear regression, adjusting for age, sex, and intracranial volume.

Sample sizes varied for each phenotype considered given missing values arising from incomplete participant questionnaires (**Supplementary Table 5**). Association P-values were FDR corrected (using the Benjamini–Hochberg procedure) to account for multiple testing within each LBC cohort.

### Variance components analysis and EWAS using BayesR+

BayesR+^31^, a software implementation of a Bayesian regression modelling framework, which implicitly controls for white cell proportions, related participants, and other unknown confounders, was used to estimate the variance accounted for in alcohol consumption by methylation data, as well as estimate its association with each measured CpG (a total of 752,722). To remove the effects of age, sex and smoking (via an EpiScore^65^), the input for BayesR+ was defined by the residuals of a linear regression model for alcohol consumption with those variables as covariates. CpG M-values were pre-corrected in a similar way, regressing out age, sex, smoking EpiScore, and batch. They were subsequently scaled to have mean zero and unit variance.

Full details of the BayesR+ modelling framework have been previously described^31^. Briefly, BayesR+ utilizes Gibbs sampling to generate draws from the posterior distribution conditional on the input data, setting prior mixture variances to 0.0001, 0.001 and 0.01, corresponding to possible small, medium and large effect sizes of the CpGs considered (explaining 0.01%, 0.1% and 1% of the variance of the phenotype of interest, respectively). After a burn-in of 5,000 samples, 10,000 samples were retained. Subsequently, a thinning of five samples was applied to reduce autocorrelation (i.e. 1,000 iterations are used when reporting results for this analysis). The convergence of the hyperparameters was evaluated through the Geweke test, as well as assessing parameter values across iterations, and assessing autocorrelation. For each probe, the proportion of iterations for which the probe was categorized as having a non-zero effect was calculated, this yielding the posterior inclusion probability (PIP). A PIP value over 0.95 (95%) was deemed to reflect an epigenome-wide significant CpG locus.

Variance components were estimated by the mean sum of squared standardised posterior effect sizes across the 1,000 iterations. Individual effect sizes were estimated as the average across the 1,000 iterations for each CpG. Models were run considering data for the full Generation Scotland cohort, as well as just the subset of “normal pattern” drinkers.

## Data Availability

All custom R (version 4.3.0), Python (version 3.9.7), and bash code is available with open access at the following GitHub repository: https://github.com/elenabernabeu/methylomics_alcohol
EWAS summary statistics will be made available on Edinburgh DataShare on publication.

## Ethics approval and consent to participate

All components of Generation Scotland received ethical approval from the NHS Tayside Committee on Medical Research Ethics (REC Reference Number: 05/S1401/89). All participants provided broad and enduring written informed consent for biomedical research. Generation Scotland has also been granted Research Tissue Bank status by the East of Scotland Research Ethics Service (REC Reference Number: 15/0040/ES), providing generic ethical approval for a wide range of uses within medical research. This study was performed in accordance with the Helsinki declaration.

Ethical approval for the LBC1921 and LBC1936 studies was obtained from the Multi-Centre Research Ethics Committee for Scotland (MREC/01/0/56) and the Lothian Research Ethics committee (LREC/1998/4/183; LREC/2003/2/29). In both studies, all participants provided written informed consent. These studies were performed in accordance with the Helsinki declaration.

Ethical approval for the ALSPAC study was obtained from the ALSPAC Ethics and Law Committee and the Local Research Ethics Committees. Consent for biological samples has been collected in accordance with the Human Tissue Act (2004). Informed consent for the use of data collected via questionnaires and clinics was obtained from participants following the recommendations of the ALSPAC Ethics and Law Committee at the time.

## Availability of data and materials

According to the terms of consent for Generation Scotland participants, access to data must be reviewed by the Generation Scotland Access Committee. Applications should be made to.

Lothian Birth Cohort data are available on request from the Lothian Birth Cohort Study, University of Edinburgh (https://www.ed.ac.uk/lothian-birth-cohorts/data-access-collaboration). Lothian Birth Cohort data are not publicly available due to them containing information that could compromise participant consent and confidentiality.

ALSPAC data are available on request from bona fide researchers. The study website contains details of all the data that is available through a fully searchable data dictionary and variable search tool (http://www.bristol.ac.uk/alspac/researchers/our-data/).

All custom R (version 4.3.0), Python (version 3.9.7), and bash code is available with open access at the following GitHub repository: https://github.com/elenabernabeu/methylomics_alcohol EWAS summary statistics will be made available on Edinburgh DataShare on publication.

## Competing interests

R.E.M has received a speaker fee from Illumina and is an advisor to the Epigenetic Clock Development Foundation. R.F.H. has received consultant fees from Illumina. R.E.M and R.F.H. have received consultant fees from Optima partners. M.R.R. receives research funding from Boehringer Ingelheim. All other authors declare no competing interests.

## Funding

### Generation Scotland

Generation Scotland received core support from the Chief Scientist Office of the Scottish Government Health Directorates (CZD/16/6) and the Scottish Funding Council (HR03006). Genotyping and DNA methylation profiling of the Generation Scotland samples was carried out by the Genetics Core Laboratory at the Edinburgh Clinical Research Facility, Edinburgh, Scotland and was funded by the Medical Research Council UK and the Wellcome Trust (Wellcome Trust Strategic Award STratifying Resilience and Depression Longitudinally (STRADL; Reference 104036/Z/14/Z). The DNA methylation data assayed for Generation Scotland was partially funded by a 2018 NARSAD Young Investigator Grant from the Brain & Behavior Research Foundation (Ref: 27404; awardee: Dr David M Howard) and by a JMAS SIM fellowship from the Royal College of Physicians of Edinburgh (Awardee: Dr Heather C Whalley).

### Lothian Birth Cohorts

We thank the LBC1921 and LBC1936 participants and team members who contributed to these studies. The LBC1921 was supported by the UK’s Biotechnology and Biological Sciences Research Council (BBSRC), The Royal Society, and The Chief Scientist Office of the Scottish Government. The LBC1936 is supported by the BBSRC, and the Economic and Social Research Council [BB/W008793/1] (which supports S.E.H.), Age UK (Disconnected Mind project), the Milton Damerel Trust, the Medical Research Council (MR/M01311/1), and the University of Edinburgh. Methylation typing of LBC1936 was supported by the Centre for Cognitive Ageing and Cognitive Epidemiology (Pilot Fund award), Age UK, The Wellcome Trust Institutional Strategic Support Fund, The University of Edinburgh, and The University of Queensland. Genotyping was funded by the BBSRC (BB/F019394/1). S.R.C. is supported by a Sir Henry Dale Fellowship jointly funded by the Wellcome Trust and the Royal Society (Grant Number 221890/Z/20/Z).

### ALSPAC

The UK Medical Research Council and Wellcome (Grant ref: 217065/Z/19/Z) and the University of Bristol provide core support for ALSPAC. This publication is the work of the authors and Matthew Suderman will serve as guarantors for the contents of this paper. A comprehensive list of grants funding is available on the ALSPAC website (http://www.bristol.ac.uk/alspac/external/documents/grant-acknowledgements.pdf). Funding for ALSPAC DNAm measurements were supported bythe Wellcome (102215/2/13/2); the University of Bristol; the UK Economic and Social Research Council (ES/N000498/1); the UK Medical Research Council (MC_UU_12013/1, MC_UU_12013/2); and the John Templeton Foundation (60828). MS and PY work within the MRC Integrative Epidemiology Unit at the University of Bristol, which is supported by the Medical Research Council (MC_UU_00011/5).

R.F.H is supported by an MRC IEU Fellowship. M.R.R. was funded by Swiss National Science Foundation Eccellenza Grant PCEGP3-181181 and by core funding from the Institute of Science and Technology Austria. E.B. and R.E.M. are supported by Alzheimer’s Society major project grant AS-PG-19b-010.

This research was funded in whole, or in part, by the Wellcome Trust (104036/Z/14/Z, 108890/Z/15/Z, 220857/Z/20/Z, and 221890/Z/20/Z). For the purpose of open access, the author has applied a CC BY public copyright licence to any Author Accepted Manuscript version arising from this submission.

## Authors’ contributions

E.B. analysed the data. A.D.C. developed the external data replication pipeline. J.K.K. and M.S. replicated results in the Sister Study and ALSPAC cohorts, respectively. D.L.M., R.F.H., J.C., MdC.V.H., S.M.M., M.E.B., J.M.W., Y.X., D.P.S., A.C., S.E.H., A.M.M., J.A.T, P.Y., S.R.C., and K.L.E. were involved in the data generation. E.B., R.E.M., and C.A.V. drafted the initial manuscript. E.B., M.R.R., C.A.V., and R.E.M. designed the study. All authors read and approved the final manuscript.

## Acknowledgements

We are grateful to all the families who took part, the general practitioners, and the Scottish School of Primary Care for their help in recruiting them and the whole GS team that includes interviewers, computer and laboratory technicians, clerical workers, research scientists, volunteers, managers, receptionists, healthcare assistants, and nurses.

ALSPAC: We are extremely grateful to all the families who took part in this study, the midwives for their help in recruiting them, and the whole ALSPAC team, which includes interviewers, computer and laboratory technicians, clerical workers, research scientists, volunteers, managers, receptionists and nurses.

## Notes

### Author Declarations

All components of Generation Scotland received ethical approval from the NHS Tayside Committee on Medical Research Ethics (REC Reference Number: 05/S1401/89). All participants provided broad and enduring written informed consent for biomedical research. Generation Scotland has also been granted Research Tissue Bank status by the East of Scotland Research Ethics Service (REC Reference Number: 15/0040/ES), providing generic ethical approval for a wide range of uses within medical research. This study was performed in accordance with the Helsinki declaration. Ethical approval for the LBC1921 and LBC1936 studies was obtained from the Multi-Centre Research Ethics Committee for Scotland (MREC/01/0/56) and the Lothian Research Ethics committee (LREC/1998/4/183; LREC/2003/2/29). In both studies, all participants provided written informed consent. These studies were performed in accordance with the Helsinki declaration. Ethical approval for the ALSPAC study was obtained from the ALSPAC Ethics and Law Committee and the Local Research Ethics Committees. Consent for biological samples has been collected in accordance with the Human Tissue Act (2004). Informed consent for the use of data collected via questionnaires and clinics was obtained from participants following the recommendations of the ALSPAC Ethics and Law Committee at the time.

