## Supplementary Figures for "Blood-based DNA methylation study of alcohol consumption"

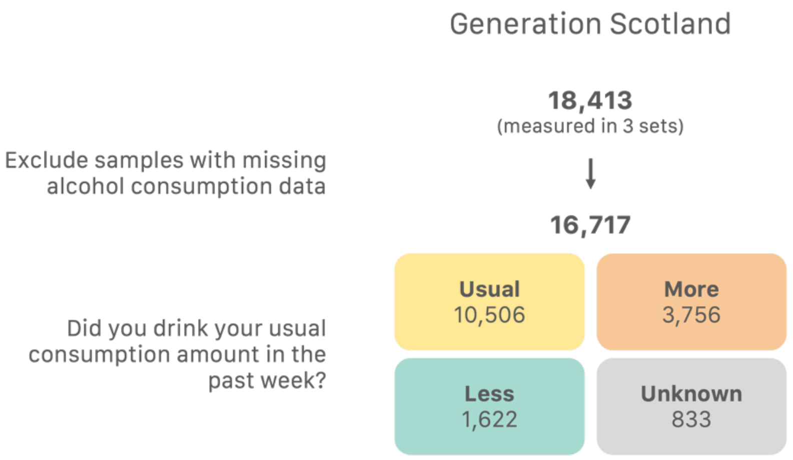


**Figure S1**. Generation Scotland sample breakdown.

a)

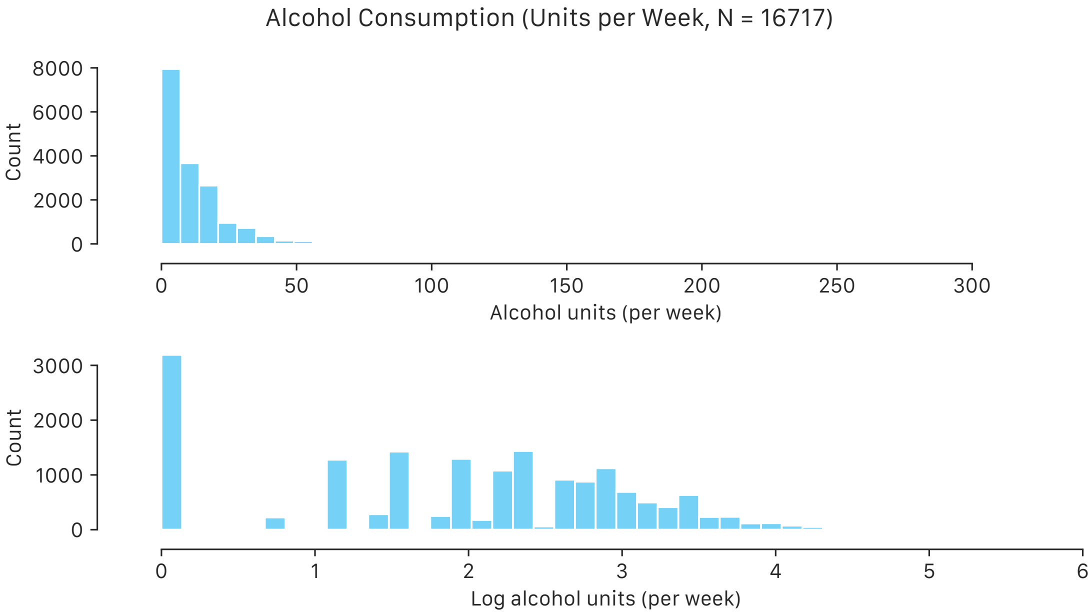


b)

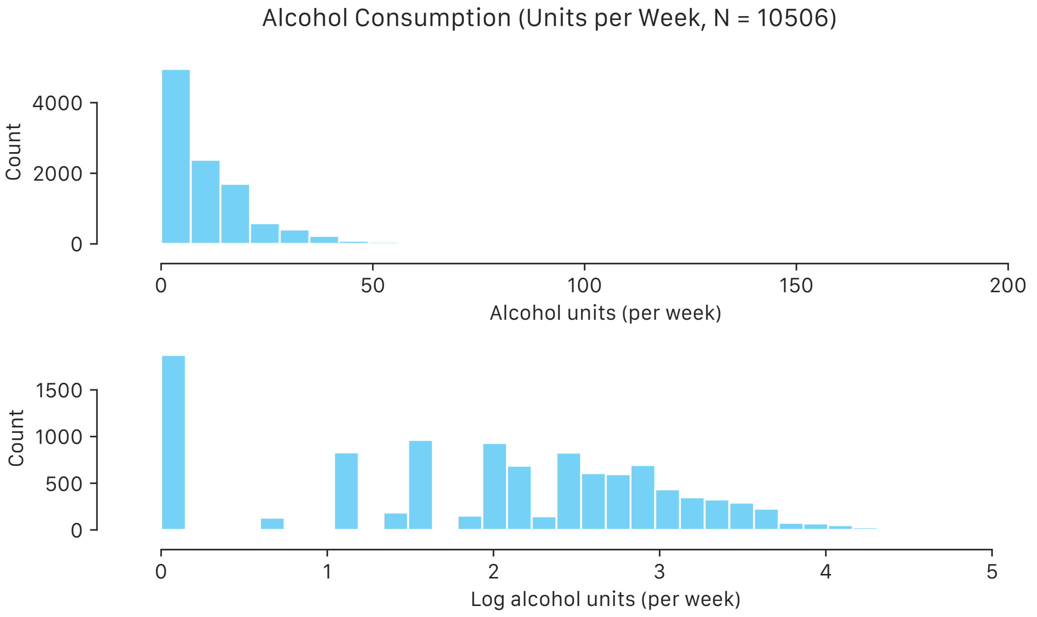


**Figure S2**. Histogram of alcohol units (measured and in log scale +1) consumed in the previous week, for a) everyone in Generation Scotland and b) just usual drinkers.

a)

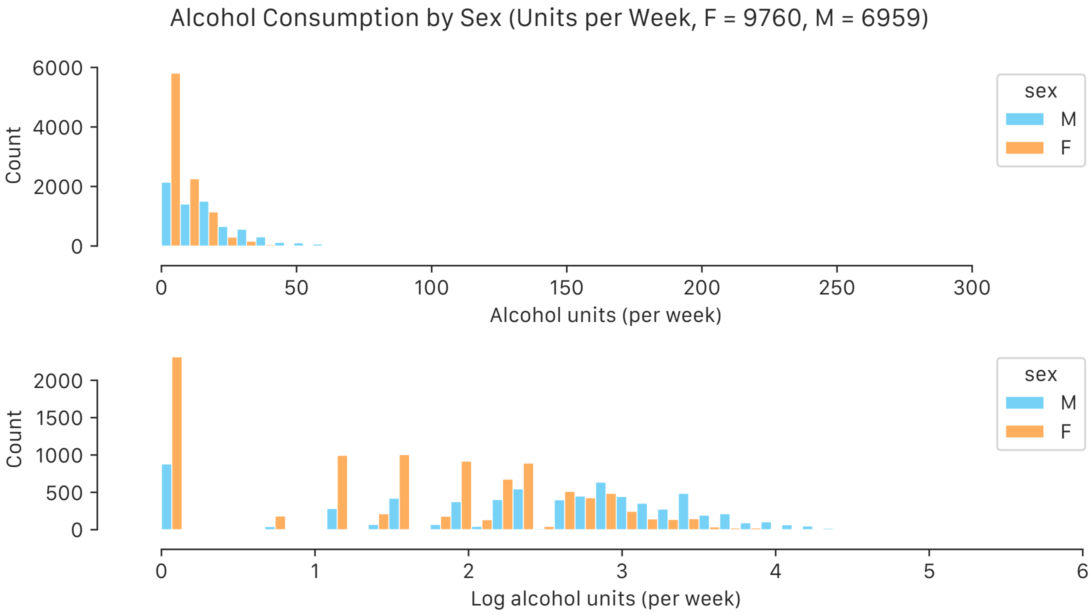


b)

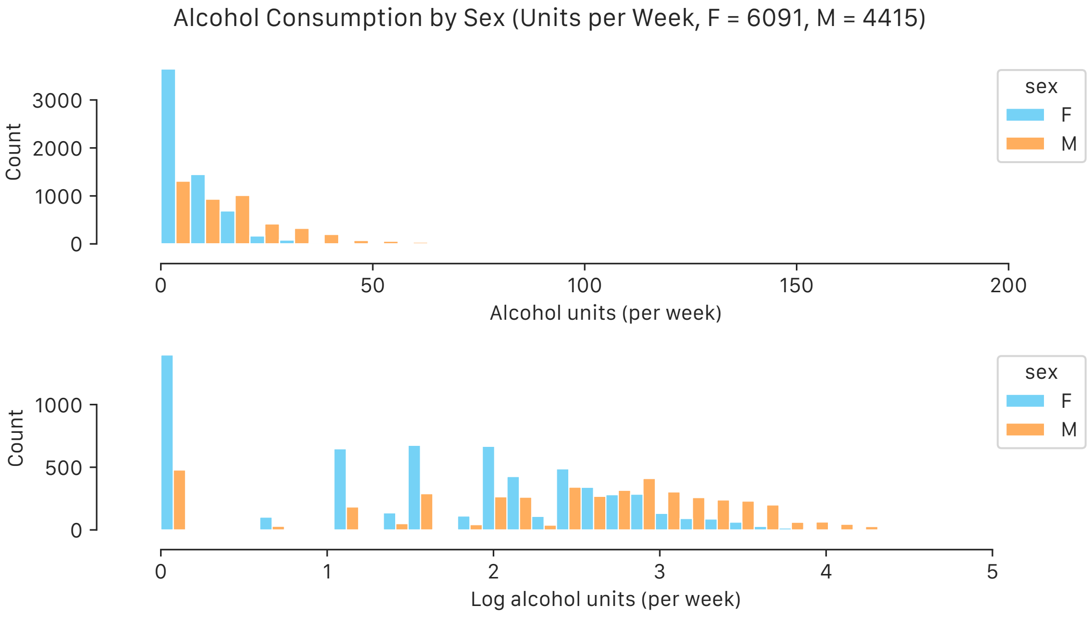


**Figure S3**. Histogram of alcohol units (measured and in log scale +1) consumed in the previous week, stratified by sex,  for a) everyone in Generation Scotland and b) just usual drinkers.

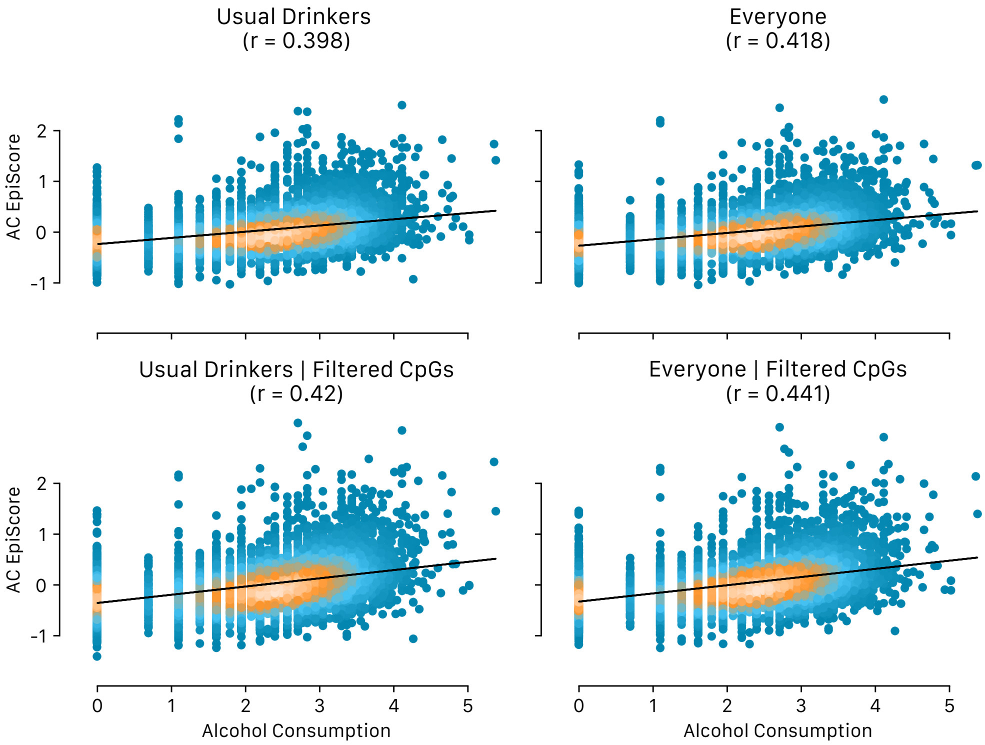


**Figure S4. Measured alcohol consumption against 4 EpiScores of alcohol consumption, trained and tested in different subsets of Generation Scotland.** Alcohol consumption (units per week) shown in log(x+1)-scale. EpiScores trained on everyone in the training set in Generation Scotland, just on usual drinkers in the training set, and filtering CpGs or not. Pearson correlations between the two measures indicated. Colour indicates density of points, with white/orange symbolising higher density.

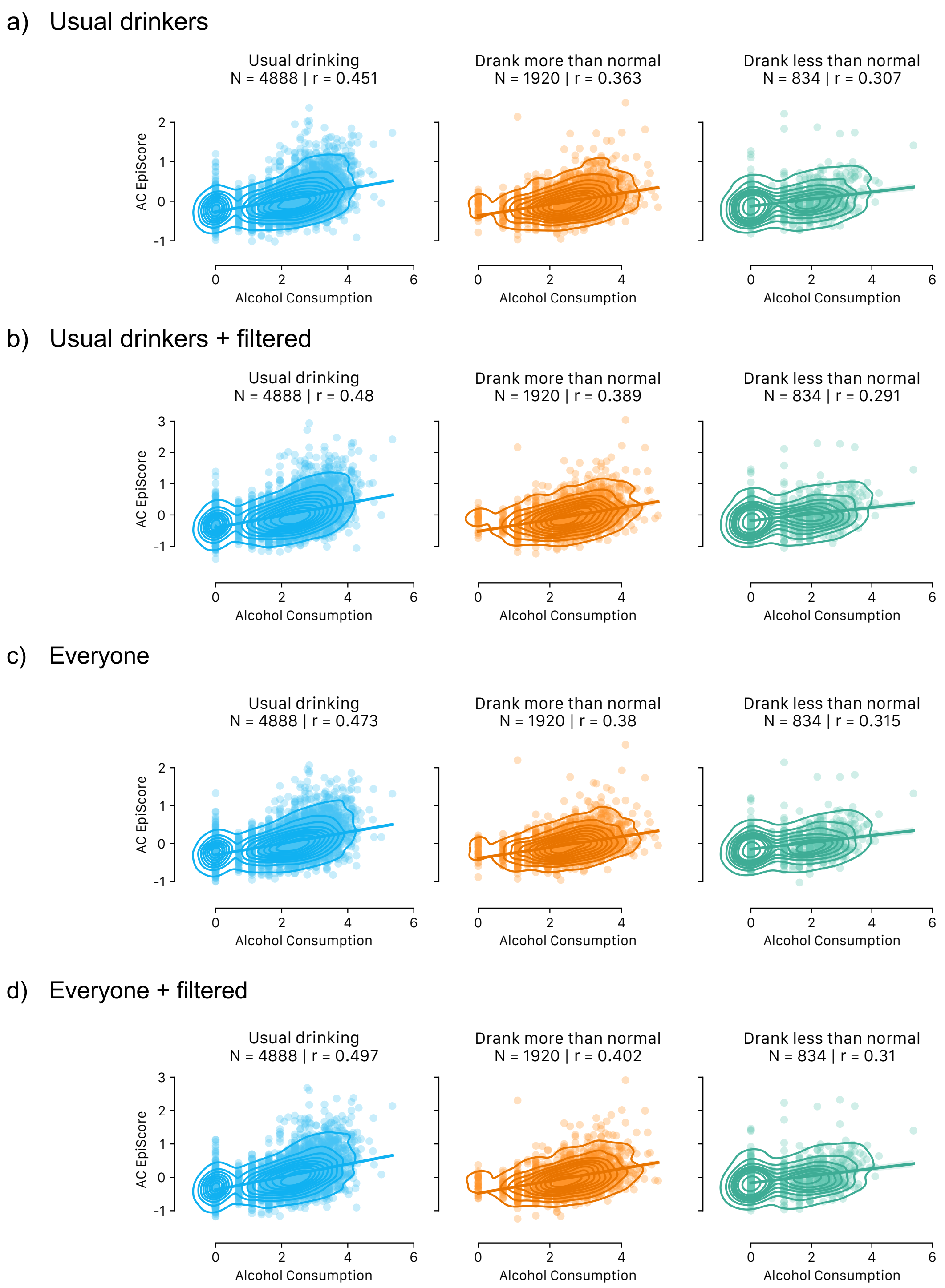


**Figure S5. Measured alcohol against 4 alcohol consumption EpiScores.** Alcohol consumption (units per week) shown in log(x+1)-scale. EpiScore trained in subset of Generation Scotland (either everyone or usual drinkers), and tested in unused subset. Prediction performance stratified by category of testing sample (usual drinker, drank more than usual in measured week, or drank less than usual in measured week). a) Trained on usual drinkers, using all CpGs, b) trained on usual drinkers using filtered CpGs, c) Trained on everyone, using all CpGs, d) trained on everyone using filtered CpGs.

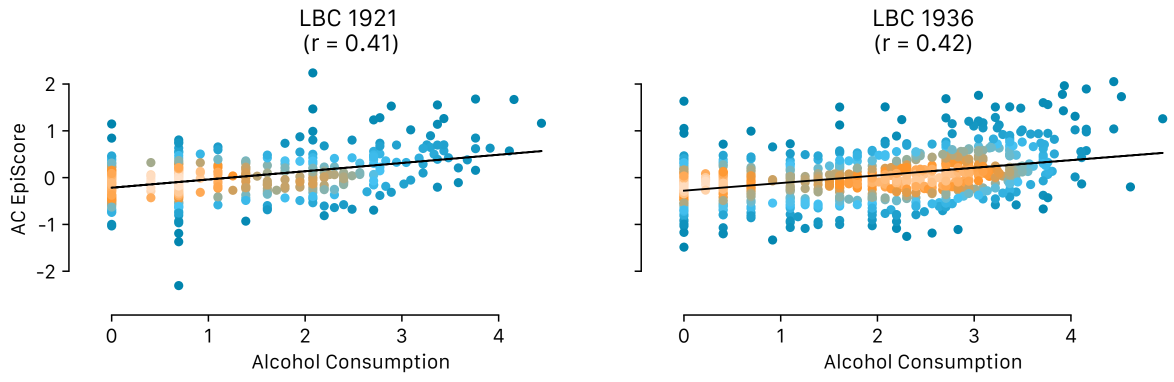


**Figure S6**. EpiScore performance in LBC1921 and LBC1936 for EpiScore trained in full Generation Scotland cohort. Alcohol consumption (units per week) in log(x+1)-scale. Colour indicates density of points, with white/orange symbolising higher density.

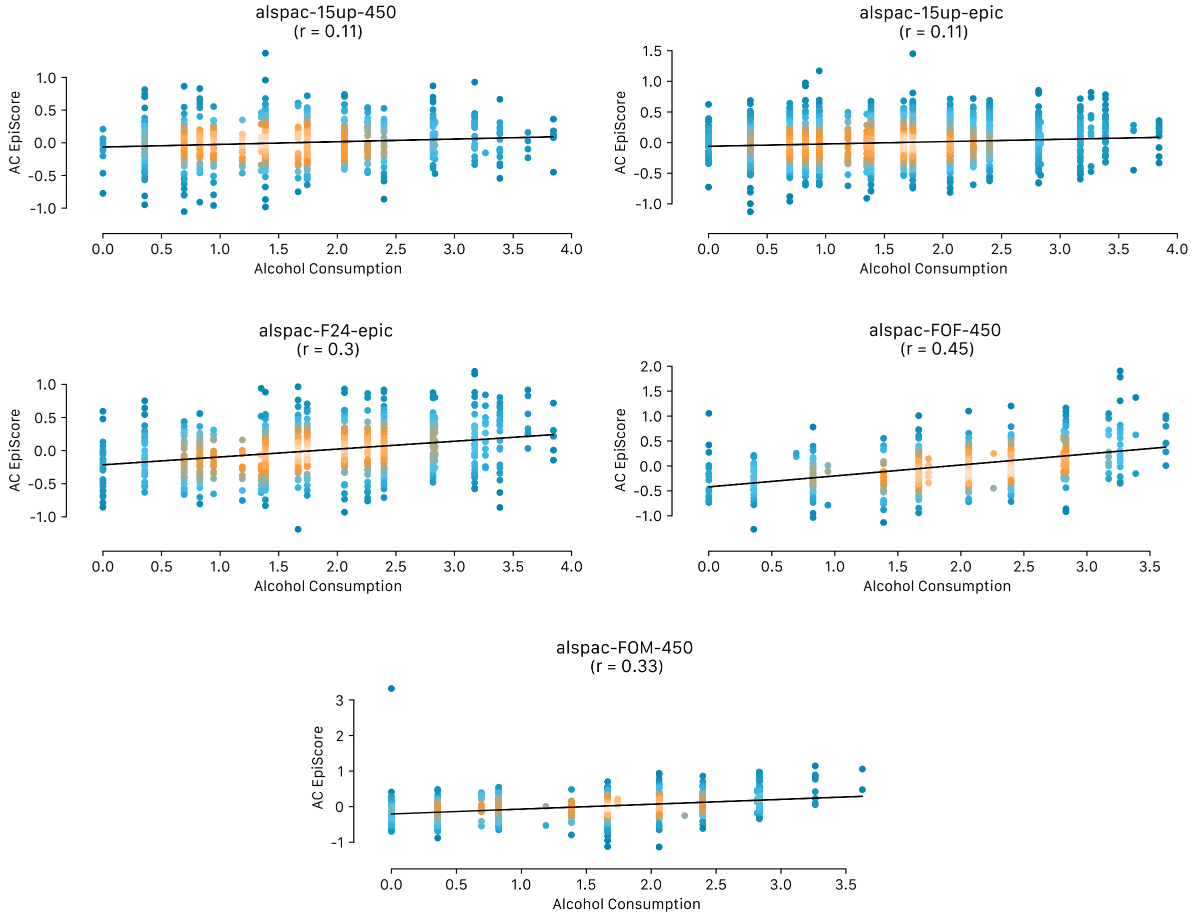


**Figure S7**. EpiScore performance across five ALSPAC cohorts. Alcohol consumption (units per week) in log(x+1)-scale. Colour indicates density of points, with white/orange symbolising higher density.

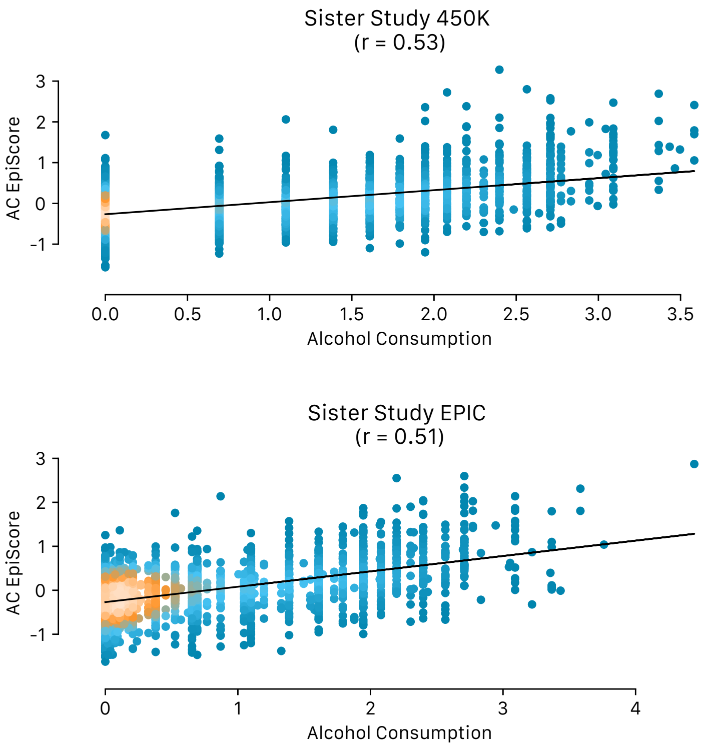


**Figure S8**. EpiScore performance across two Sister Study cohorts. Alcohol consumption (units per week) in log(x+1)-scale. Colour indicates density of points, with white/orange symbolising higher density.

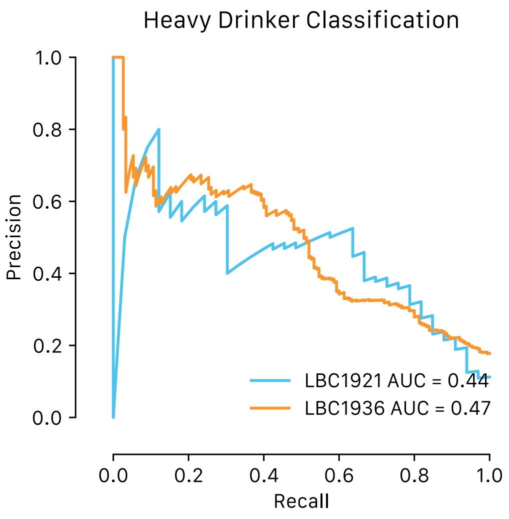


**Figure S9**. **AUC analysis of alcohol consumption prediction in the Lothian Birth Cohorts of 1921 and 1936.** Shown are area under the precision recall curves for classifying dichotomized alcohol consumption (heavy drinkers - >14 units per week for females or >21 units per week for males - versus non- or light-moderate drinkers).

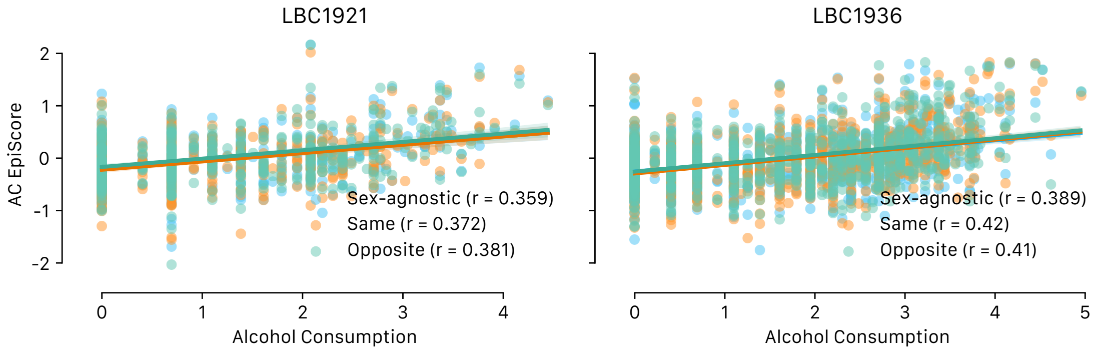


**Figure S10. Sex-specific EpiScore performance test in the Lothian Birth Cohorts.** Measured alcohol consumption (units per week) is compared against three different EpiScore modalities (sex-agnostic, same-sex, and opposite-sex) in the Lothian Birth Cohorts. Alcohol consumption in log(x+1)-scale.
